## Appendix 1: Data Collection Form for "Derivation and validation of a clinical severity score for acutely ill adults with suspected COVID-19: The PRIEST observational cohort study"

AFFIX PATIENT DETAILS LABEL HERE IF AVAILABLE

Name ☐ Male ☐ Female

Date of birth ☐☐:☐☐:☐☐☐☐

NHS Number ☐☐☐☐☐☐☐☐☐☐

Hospital Number ☐☐☐☐☐☐☐☐☐☐

**DATE:** ☐☐:☐☐:☐☐☐☐

**TIME:** ☐☐:☐☐

**PRESENTING FEATURES:**

☐ Shortness of breath ☐ Confusion ☐ Vomiting

☐ Cough ☐ Rash

☐ Fever ☐ Anosmia

☐ Sore throat ☐ Abdominal pain

☐ Headache ☐ Diarrhoea

**REFERRAL SOURCE**

☐ GP

☐ Self

☐ 111

☐ 999

☐ Other

**PREVIOUS** ☐ Flu Vaccine?[1] ☐ Oseltamivir?[2] ☐ Previous Attendance?[3]

**ANTIBIOTIC THERAPY THIS ILLNESS?** ☐ None
*(Drug and duration)*

**SYMPTOM DURATION**
(days)

**CURRENT MEDICATION** ☐ None

**ALLERGIES TO MEDICATION** ☐ None

**MEDICAL HISTORY / CHRONIC DISEASE** ☐ None

☐ Heart disease ☐ Diabetes ☐ Hypertension

☐ Renal impairment ☐ Active malignancy *(last 6 months)*

☐ Steroid therapy ☐ Immunosuppression

☐ Asthma ☐ Other chronic lung disease

**RECENT TRAVEL HISTORY –** last two weeks
*(Country, duration and days since return)*

**LIFESTYLE** ☐ Patient lives alone / no fixed abode ☐ Clinically obese ☐ Pregnant

☐ Tobacco user ☐ Vape user

☐ Known contact with Covid-19 case

*Relationship to contact*

**PAEDIATRIC ONLY**

☐ Routine vaccinations? ☐ Taking feeds ☐ Parental anxiety? ☐ Premature[4]

**PERFORMANCE STATUS** (***tick one***)

☐ Unrestricted normal activity ☐ Limited strenuous activity, can do light ☐ Limited activity, can self care

☐ Limited self care ☐ Bed/chair bound, no self care

**WITHDRAWN CASE?** Leaflet ☐ Verbal ☐

### PANDEMIC RESPIRATORY INFECTION FORM

[1]Yes if any previous vaccine [2]Yes if any use of oseltamivir in current illness
[3]Yes if previous attendance at emergency dept. for this problem

[4]Premature defined as birth before 37 weeks gestation.

Version 7.0 27th Apr 2020

#### CLINICAL EXAMINATION

***MOST LIKELY DIAGNOSIS?*** ☐ Influenza (Pandemic or seasonal) ☐ Covid-19 ☐ *Other (provide details)*

| | |
|---|---|
| Respiratory Rate | |
| Pulse Rate | |
| Temperature | |
| Blood Pressure | |
| $SaO_2$ | |
| Post exercise SaO2 *(if measured)* | |
| $FiO_2$ | |
| Central capillary refill | Normal ☐ Abnormal ☐ |
| GCS Total | ☐ GCS individual scores not available |

| GCS-E | GCS-V | GCS-M |
|---|---|---|

| A | V | P | U |
|---|---|---|---|

☐ Severe respiratory distress[1] ☐ Respiratory exhaustion ☐ Severe dehydration

#### OTHER CLINICAL CONCERNS:

| | | | |
|---|---|---|---|
| CXR | Not done ☐ | Normal ☐ | Abnormal ☐ |
| ECG | Not done ☐ | Normal ☐ | Abnormal ☐ |

**BLOODS TAKEN** ☐

| Na | K | Urea | Creat | Hb | Plate | WCC | Lymp | Neut | Lac - tate | CRP | D-dimer | Trop - onin |
|---|---|---|---|---|---|---|---|---|---|---|---|---|

#### DISPOSITION AND CLINICAL PLAN

*Oseltamivir* ☐ *Antibiotic* [ ]

*Antibiotic details*

Clinician Name: Signature: Grade:

Disposed to: Date: Time:

### PANDEMIC RESPIRATORY INFECTION FORM

[1]Severe respiratory distress (accessory muscles, tracheal tug, feeling of suffocation, apnoea)
