## Appendix 2: Follow-up Form for "Derivation and validation of a clinical severity score for acutely ill adults with suspected COVID-19: The PRIEST observational cohort study"

AFFIX PATIENT DETAILS LABEL HERE IF AVAILABLE

NHS Number ☐☐☐☐☐☐☐☐☐☐

Hospital Number ☐☐☐☐☐☐☐☐☐☐

**DATE OF INITIAL VISIT:** ☐☐:☐☐:☐☐☐☐

**30 DAYS DUE AT:** ☐☐:☐☐:☐☐☐☐

**Mortality status**

☐ Alive ☐ Dead → Date of death ☐☐ ☐☐ ☐☐☐☐ (d d m m y y y y)

**Respiratory pathogen confirmed** ☐ Influenza (Pandemic or seasonal)

☐ Covid-19 ☐ Other *(provide details)*

Date pathogen confirmed ☐☐ ☐☐ ☐☐☐☐ (d d m m y y y y)

**Was patient admitted at initial assessment**

☐ Yes ☐ No → ☐ Discharged home ☐ Transferred to other hospital ☐ Other / destination unknown

**Admissions up to 30 days:** *using patient's notes please complete as fully as possible, indicating dates for events and what support provided. Please include initial assessment if patient was admitted.*

| Reason for admission | Ward | Admission dates | Respiratory support | | Cardiovascular support | | Renal support | |
|---|---|---|---|---|---|---|---|---|
| | | | Days | Type | Days | Type | Days | Type |
| | ☐ Ward<br>☐ ITU<br>☐ HDU | Start ☐☐ ☐☐ ☐☐☐☐ (d d m m y y y y)<br>End ☐☐ ☐☐ ☐☐☐☐ (d d m m y y y y) | | | | | | |
| | ☐ Ward<br>☐ ITU<br>☐ HDU | Start ☐☐ ☐☐ ☐☐☐☐ (d d m m y y y y)<br>End ☐☐ ☐☐ ☐☐☐☐ (d d m m y y y y) | | | | | | |
| | ☐ Ward<br>☐ ITU<br>☐ HDU | Start ☐☐ ☐☐ ☐☐☐☐ (d d m m y y y y)<br>End ☐☐ ☐☐ ☐☐☐☐ (d d m m y y y y) | | | | | | |

**<u>30 day follow-up – for researcher use only</u>**

AFFIX PATIENT DETAILS LABEL HERE IF AVAILABLE

NHS Number ☐☐☐☐☐☐☐☐☐☐

Hospital Number ☐☐☐☐☐☐☐☐☐☐

Was a DNR decision made at any time between initial presentation and follow-up ☐ Yes ☐ No → Date ☐☐ ☐☐ ☐☐☐☐ (d d m m y y y y)

**If patient experienced any events that did not require respiratory, cardiovascular or renal support, but that:**

- **were life threatening,**
- **resulted in persistent or significant disability or incapacity**
- **prolonged hospitalisation**

**Please add details below**

**Adverse events:** using patient's *notes please complete as fully as possible, indicating dates for events and what support provided*

| Event details | Event dates | Seriousness |
|---|---|---|
| | Start ☐☐ ☐☐ ☐☐☐☐ (d d m m y y y y)<br>End ☐☐ ☐☐ ☐☐☐☐ (d d m m y y y y) OR ☐ Ongoing | ☐ Life threatening<br>☐ Persistent or significant disability or incapacity<br>☐ Prolongs hospitalisation |
| | Start ☐☐ ☐☐ ☐☐☐☐ (d d m m y y y y)<br>End ☐☐ ☐☐ ☐☐☐☐ (d d m m y y y y) OR ☐ Ongoing | ☐ Life threatening<br>☐ Persistent or significant disability or incapacity<br>☐ Prolongs hospitalisation |
| | Start ☐☐ ☐☐ ☐☐☐☐ (d d m m y y y y)<br>End ☐☐ ☐☐ ☐☐☐☐ (d d m m y y y y) OR ☐ Ongoing | ☐ Life threatening<br>☐ Persistent or significant disability or incapacity<br>☐ Prolongs hospitalisation |

**30 day follow-up – for researcher use only**
