## Appendix 3: Study steering committee for "Derivation and validation of a clinical severity score for acutely ill adults with suspected COVID-19: The PRIEST observational cohort study"

| **Title** | **First Name** | **Last Name** | **Job Title** | **Name of employing institution, and any institutions where this nominee holds an Honorary Contract** | **Membership Type:** | **Independent** | **Expertise** |
| --- | --- | --- | --- | --- | --- | --- | --- |
| Mrs | Shan | Bennett | PPI |  | PPI Member | Yes | PPI, Sheffield Emergency Care Forum |
| Prof  (Associate Professor) | Paul | Baxter | Senior Lecturer in Biostatistics | University of Leeds.  Honorary contract with Leeds Teaching Hospitals NHS Trust. | Member | Yes | Biostatistics |
| Prof | Tim | Coats | Professor of Emergency Medicine | University of Leicester | Chair | Yes | Clinician Emergency Medicine Research |
| Mrs | Enid | Hirst | Co-ordinator of Sheffield Emergency Care Forum (PPI) |  | PPI Member | Yes | PPI, Sheffield Emergency Care Forum |
| Mrs | Beryl | Darlison | PPI |  | PPI Member | Yes | PPI, Sheffield Emergency Care Forum |
| Dr | Kavin | Smith | Deputy Director Healthcare Public Health England, Yorkshire and the Humber (Replaced Will Morton as the PHE representative) | Public Health England | Member | Yes | Public health |
| Dr | Will | Morton | Consultant in Health Protection at Public Health England | Public Health England. Honorary contract with the University of Manchester. | Member | Yes | Health protection specialist |
| Dr | Nazir | Lone | Senior Clinical Lecturer in Critical Care | The University of Edinburgh, Honorary Consultant in Critical Care at the Royal Infirmary of Edinburgh. | Member | No | Cliniciain Critical Care, Critical Care Epidemiology |
| Dr | Graham | McClelland | Research paramedic | North East Ambulance Service NHS Trust | Member | Yes | Clinician Paramedic |
| Prof | Steve | Goodacre | PRIEST CI | The University of Sheffield | Member | No | Clinician Emergency Medicine Research |
| Mrs | Rachel | Robinson | Chief Nurse | Integrated Care 24 Ltd | Member | Yes | Clinician, 111 Knowledge |
| Dr | Mathew | Beattie | Medical Director North East Ambulance Service Foundation Trust | North East Ambulance Service Foundation Trust | Member | Yes | Clinician, North East Ambulance Service |
