## Appendix 4: Site research staff for "Derivation and validation of a clinical severity score for acutely ill adults with suspected COVID-19: The PRIEST observational cohort study"

Anna Wilson

Bethan Holroyd-Hind

Chris Fitzsimmons

Julie Morcombe

Arianna Bellini

Chloe Lyons

Tracy Marsden

Mr Paul Brittain

Mrs Claire Brookes

Mrs Heidi Redfearn

Dr Ben Bloom

Imogen Skene

Raine Astin-Chamberlain

Laura Barman

Janine Mallison

Lisa Baldwin

Anneli James

Lucy Newton

Mark D Lyttle

Rebecca Hoskins

Sally Melson

Bristol Clinical Trials Team

Mrs Emma Storr

Mr Martin Walton

Dr Felix Wood

Annie Rose RGN

Francesca Wright RGN

Aarzoo Khan

Liza Keating

Emma Craig

Elizabeth Taylor

Andrew Rees MBA RICR

Simon Sharpe

Heather Sellers

Dr Tanya de Weymarn

Gloucestershire Cancer Clinical Trials Team

Jagtar Pooni

Sara Simmons

Dani Steward

Dr Adrian Marsh

Sr Mandy Carnahan

Sr Lucy Price

Dr Mark Harrison

Rebecca Emmonds

Jane Luke

Northumbria Clinical Trials Team

Mrs Katrina Parkinson

Miss Georgia Thomasson

Mrs Alda Remegoso

Bernard Hadebe

Joanne Galliford

Prisca Gondo

Paula Harman

Melanie Darwent

Ross Downes

Sally Beer

Dr Jennifer Lockwood

Harrogate Research team

Alasdair Gray

Polly Black

Amanda Lyle

Yvonne Lester

Dane Goodere-Bennett

Dr Huw Steven Jenkins

Dr John Wright

Kimberley Webster

Professor Richard Body

Dr Eloise Cook

Non-clinical COVID-19 Research Delivery Team at Manchester

Abdo Sattout

Melanie Harrison

Sarah Stevenson

Dr Adrian Boyle

Susie Hardwick

Debbie Read

Frank Coffey

Megan Meredith

Helen Navarra

Mrs Judith Ratcliff

Fiona Thompson

Amanda Adamson

Dr Gareth Hampton

Dr Sarah Wilson

Mrs Joana Da Rocha

Dr Charlotte Griffiths

Dr Nam Tong

Mrs Tracy Fuller

Mrs Hannah Bloxham

Alastair Richards

Debra Barnett

Lindianne Aitken

Suzannah Pegler

Maggie Walton

Tim Slade

Fleur Cantle

Hannah Cotton

Maeve Cockrell

Jessica Law

Ava Williams

Janet Mills

Janice Birt

Cassandra Gleeson

Dr Recebba Macfarlane

Mrs Lisa Evans

Ms Eloise Van Vuren

Dr Amelia Gruber

Dr Ignacio Cardona

Laura O'Rourke

Julie Quigley

Mohammad Zubair Ahmad

Daniella Hydes

Suzanne Mason

Mishel Cunningham

Nicola Lancaster

Amanda Cowton

Sarah Clark

Jane Varin

Karl Ward

Ella Sykes

Heather Jarman

Desislava Baramova

Marta Pizzorusso

Dr Sarah Essex

Mrs Andrea Watson

Mr Craig Mower

Sara Bennett

Judith Bell

Abigail Pemberton

Dr Jill Woodhead

Sherwood Forest Clinical Trials Team

Dr Amber Nocher

Dr Henrietta Morton King

Mrs Jo-Ann Taylor

Dr Shayma Habeeb

Wojciech Sawicki

Kate Martin

Nicola Charnley

Mr Matthew Edward Ryan

Dr Shrouk Messahel

Dr Daniel B Hawcutt

Miss Laura Purandare

Mr Daniel Griffiths

Miss Rebecca Miln

Robert Hull

Laura Robertson

Michaela Sutherland

Bolton Clinical Trials Team

Christine Dixon

Ellen Jessup-Dunton

Reina Layug

Dr. Rajendar Garlapati

Farzana Masters

Yvonne Grimes

Joseph Dykes

Katharine Gantert

Favour Chukwunonyerem

Dilara Arslan
