## Appendix 5: CTRU acknowledgements for "Derivation and validation of a clinical severity score for acutely ill adults with suspected COVID-19: The PRIEST observational cohort study"

**Appendix 5: Supporting Research Staff**

Marie Hyslop

Dan Beever

Samuel Keating

Kerry Wilson

Heather Dakin

Edwin Burkinshaw

Kirsty Pemberton

Tim Chater

Chris Turtle

Emily Turton

Matthew Bursnall

Mike Bradburn

Jennifer Petrie

Lizzie Swaby

Gemma Hackney

Judith Cohen
